# Examining spousal and family support as predictors of long-term weight loss and remission of type 2 diabetes following bariatric surgery

**DOI:** 10.1101/2021.06.27.21259510

**Authors:** Edina YQ Tan, Phong Ching Lee, Kwang Wei Tham, Sonali Ganguly, Chin Hong Lim, Jean CJ Liu

## Abstract

**Background:** Postoperative outcomes vary considerably across bariatric patients and may be related to psychosocial factors. In this study, we examined whether a patient’s family support predicts weight loss and the remission of type 2 diabetes (T2DM) up to 5 years following surgery.

**Methods:** Between 2008 to 2018, 359 patients completed a pre-surgical questionnaire before undergoing gastric bypass or sleeve gastrectomy. As part of the questionnaire, patients described their family support in terms of structure (marital status, number of family members in the household) and function (marriage satisfaction, family emotional support, family practical support). These were applied as predictors to percent total weight loss (%TWL) and T2DM remission at 1, 2, 3, 4, and 5 years following surgery.

**Results:** Marital satisfaction was found to be a significant predictor of post-surgical weight trajectories. Namely, patients who reported higher marital satisfaction were more likely to sustain weight loss than patients who reported lower marital satisfaction (β = 0.92, SE = 0.37, *p* = 0.02).

**Conclusions:** Given the link between marital support and long-term weight outcomes, providers could consider asking patients about their spousal relationships during pre-surgical counselling.

**Key Points:**

⍰ Psychosocial factors may influence postoperative outcomes
⍰ Family support was examined as a predictor of weight loss and type 2 diabetes
⍰ Marital satisfaction significantly predicted post-surgical weight trajectories

## Introduction

Although bariatric surgery produces marked weight loss and decreased obesity-related comorbidities (1), post-surgical outcomes vary across individuals. Body mass index (BMI) decreases by an average of 13 kg/m^2^ in the first year (1), but weight trajectories differ: whereas some patients maintain the initial weight loss or persist in further weight loss, others regain the weight over time (2). Similarly, individuals differ in whether they experience remission of diabetes mellitus, hypertension, and dyslipidemia, and whether these changes are sustained (2). Given these individual differences, there is a need to identify factors that can predict postoperative outcomes, allowing patient selection and follow-up care to be optimised.

One potential predictor may be a patient’s family support. Within the hospital setting, family support has been linked to a wide range of patient outcomes, including: duration of stay in intensive care units (3), management of chronic illnesses (4), and quality of life for paediatric patients after open-heart surgery (5). In the case of bariatric surgery, the impact of family support is less clear (6).

Family support can be defined either by its structure or by the function it serves (7). Structural support refers to the availability of family members, based – for example – on marital status or the number of family members living within the household (7). Using this definition, several studies have described how married patients experience less weight loss than single patients 12- to 24-months following surgery; however, other studies either did not find this association, or reported the inverse relationship (8-10). Beyond these mixed findings, little is known about the longer-term impact of marital status, the impact of having non-spousal family members, or the impact of family structure on comorbidities (8).

Aside from structure, family support can also be described by function – the extent to which patients receive emotional or practical support from family members (7). When interviewed, patients who succeed in weight maintenance typically recount how family members encouraged or assisted them with everyday tasks (11, 12); conversely, those who experienced weight regain reported feeling lonely (13). Despite these accounts, we are aware of only five studies that have quantified the impact of functional support. Two studies reported increased weight loss amongst patients who received dietary and lifestyle support (14), or who reported better family functioning (including better emotional support) (15); however, a third study found no impact of family attendance at medical appointments (16). Specific to spouses, two studies reported that marriage satisfaction – a proxy marker for the quality of spousal support received – predicted weight status at 6 and 12 months post-surgery (17, 18).

To summarise, a limited number of quantitative studies have examined whether family support predicts long-term outcomes after bariatric surgery. Preliminary findings suggest that the nature and quality of family support (functional support) may be better predictors than the number of family members available (structural support). However, this hypothesis has not been tested directly. Further, prior research has almost exclusively focused on weight loss as the outcome measure, and has examined brief postoperative time windows (6-36 months). Correspondingly, little is known about the remission of comorbidities, or the longer-term impact of family support on weight trajectories.

To address these gaps, we characterized preoperative structural and functional family support in a cohort of patients undergoing bariatric surgery. As our primary goal, we sought to identify which aspect of family support would predict weight loss and complete remission of type 2 diabetes (T2DM) up to five years following surgery.

## Methods

### Study Design and Population

We conducted a chart review of patients who had undergone laparoscopic sleeve gastrectomy (LSG), one anastomosis gastric bypass (OAGB) or Roux-en-Y gastric bypass (RYGB) at a Singapore-based tertiary centre from 2008 to 2018 (19). All patients met local criteria for surgery (BMI ≥ 37.5kg/m^2^, or BMI ≥ 32.5kg/m^2^ with comorbidities), and were included if they had completed an intake questionnaire on their medical and social history (routinely administered to patients during their first pre-operative visit). Data were retrieved from electronic medical records managed using the Research Electronic Data Capture platform (REDCap). The study was approved by the SingHealth Centralized Institutional Review Board (2011/054/C), and was registered on ClinicalTrials.gov (NCT04303611).

### Outcome measures

#### Weight loss outcomes

Weight in kilograms was measured pre-surgery (on the day of surgery), and at follow-up clinical appointments 1, 2, 3, 4, and 5 years post-surgery. This was expressed as total percentage weight loss (%TWL), defined as the difference between patients’ post-surgical weight (at the follow-up appointment) and pre-surgical weight, divided by pre-surgical weight and expressed as a percentage.

#### Complete T2DM remission

Similarly, percentage of glycated hemoglobin (HbA1c) was measured pre-surgery and at clinical appointments 1, 2, 3, 4, and 5 years post-surgery. Complete T2DM remission was defined as HbA1c < 6.0% without the use of anti-diabetic medication.

### Covariates

#### Spousal support

Spousal support was first defined structurally, based on patients’ self-reported marital status on the intake questionnaire (married, not married: single/separated/ divorced/widowed). As a marker for functional support, patients also rated their marital satisfaction using a 4-point scale ranging from “Dissatisfied” (1) to “Very Satisfied” (4). (This variable was left empty for patients who were not married.)

#### Family support

Within the intake questionnaire, patients also listed the name, age, and relationship of every person living in their household. Structural family support was then derived by summing up the total number of family members listed (≤ 2 or > 2).

Two further questions assessed functional support. For each family member listed, participants rated whether the person supported their weight loss efforts (1: yes, 0: not involved, -1: no). Scores were then summed to provide a measure of emotional support, with total scores recategorized as: ‘unsupportive’ (≤ -3 to -1), ‘neutral’ (0) or ‘supportive’ (1 to ≥ 3).

As an assay of practical support, patients indicated the primary person within the household who: (i) planned meals, (ii) cooked, and/or (iii) visited the supermarket or wet market for groceries. Based on their responses, patients were then classified as ‘having practical support’ (if at least one family member either planned meals, cooked, or purchased groceries), or ‘having no practical support’ (if no family member performed these roles).

#### Demographic and clinical variables

As additional covariates, we extracted: gender (male, female), ethnicity (Chinese, Malay, Indian, Others), age at time of surgery, BMI at time of surgery, and type of surgery (LSG, OAGB, RYGB).

### Statistical Analyses

Statistical analyses were conducted using SPSS 25 (IBM Corp., Armonk, NY) and R 3.4.0 (R Core Team, Vienna, Austria). To track changes in %TWL over time, we ran a linear mixed-effects model with parameters estimated using maximum likelihood estimation (using a first-order autoregressive covariance structure). We included as fixed effects: time (years following surgery), the five family support variables (marital status, marital satisfaction, number of family members, emotional family support, and structural family support), and the interaction between time and the family support variables [Model 1]. Random intercepts accounted for correlated data due to the longitudinal design, and continuous variables were grand mean-centered. To assess robustness, we repeated this analysis while controlling for the set of demographic and clinical variables [Model 2].

For T2DM remission, we ran two Cox proportional-hazard models to investigate the association between covariates and the time taken for complete remission to occur. As with %TWL, Model 1 included the five family support variables, while Model 2 further controlled for demographic and clinical variables.

## Results

359 patients were included in the analyses (Table 1). 62.7% were female, with a mean preoperative age of 40.1 ± 10.1 years and BMI of 42.6 ± 7.7 kg/m^2^. The majority underwent LSG (75.2%). At baseline, 37.3% of the sample had T2DM with a mean preoperative HbA1c (%) of 6.82 ± 1.67.

As shown in Figure 1, mean %TWL following surgery was: 27.4 ± 7.9 (1 year post-surgery), 26.3 ± 8.7 (2 years), 24.9 ± 8.34 (3 years), 22.9 ± 10.0 (4 years), and 22.1 ± 9.84 (5 years). Mean postoperative HbA1c (%) was: 5.44 ± 0.7 (1 year post-surgery), 5.90 ± 1.2 (2 years), 6.06 ± 1.2 (3 years), 6.2 ± 1.2 (4 years), and 6.2 ± 1.2 (5 years).

**Figure 1.**
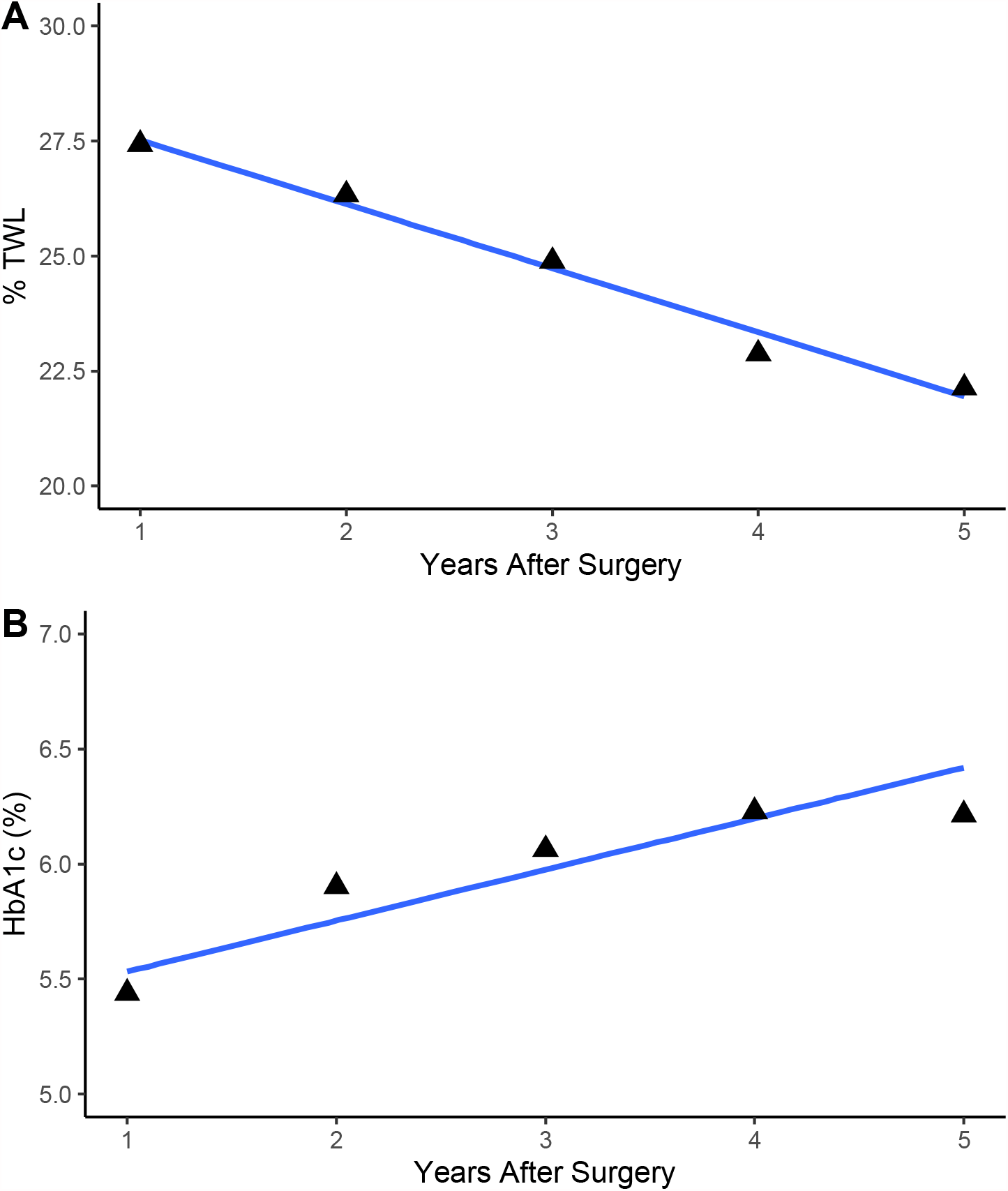
Changes in: (A) percent total weight loss (%TWL), and (B) glycated hemoglobin (HbA1c) up to 5 years following bariatric surgery. Grey bands represent the 95% confidence intervals.

### Presurgical Family Support

#### Spousal support

57.9% of patients were married at the time of surgery, with most reporting marital satisfaction (78.3% satisfied or very satisfied; Table 1).

#### Family support

36.2% of patients had more than 2 family members living in the same household. Most patients (73.5%) reported having practical family support, with at least one family member assisting with meal-planning, cooking, or groceries. Similarly, 73.3% of patients reported having emotional support, with family members supportive of their weight loss. Where patients had unsupportive family members, these tended to be their parents (47.8%).

### Predictors of Postoperative %TWL

As shown in Table 2, marital satisfaction significantly predicted the trajectory of %TWL over 5 years (Model 1: β = 0.92, SE = 0.37, *p* = 0.02). Namely, patients who reported higher preoperative marital satisfaction showed more prolonged maintenance of weight loss post-surgery (i.e., less steep declines), compared to patients who reported lower marital satisfaction (Figure 2). This effect was robust, and was observed even when demographic and clinical variables were controlled for (Model 2: β = 0.79, SE = 0.37, *p* = 0.04).

**Figure 2.**
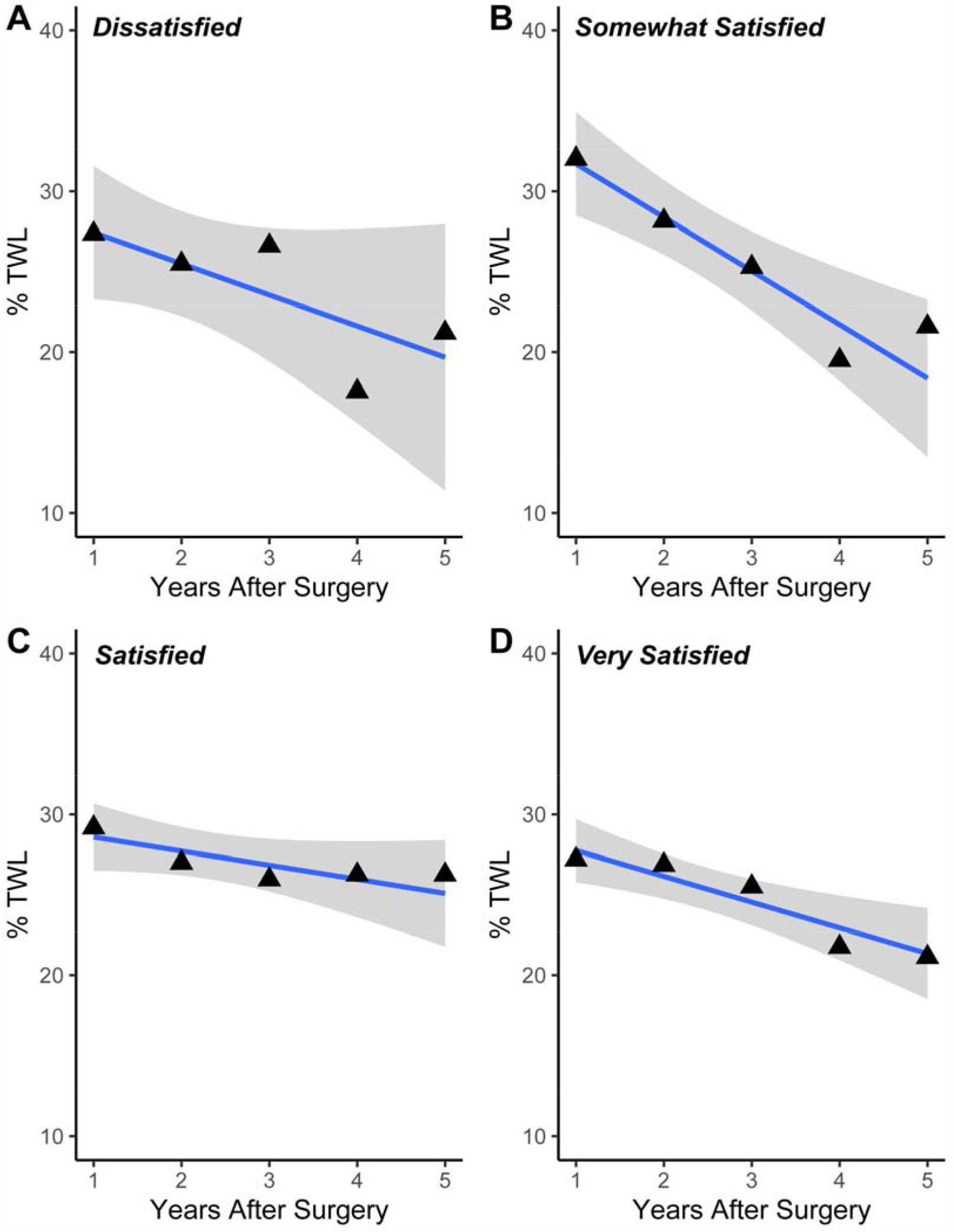
Weight trajectories as a function of preoperative marital satisfaction (A: dissatisfied, B: somewhat satisfied, C: satisfied, D: very satisfied). Patients with higher levels of marriage satisfaction showed more prolonged maintenance of percent total weight loss (%TWL), as evidenced by less steep declines in %TWL. Grey bands represent the 95% confidence intervals.

Averaged across time, preoperative BMI (β = 0.51, SE = 0.11, *p* < 0.001) and type of surgery (RYGB: β = 2.18, SE = 0.92, *p* = 0.02) also emerged as predictors of %TWL. No other main or interaction effect was significant (smallest *p* = 0.06).

### Predictors of T2DM Control

For T2DM management, preoperative age (HR = 0.97 [95% CI: 0.95, 0.99], *p* = 0.01), preoperative BMI (HR = 1.03 [95% CI: 1.00, 1.06], *p* = 0.02), and type of surgery significantly predicted complete remission (OAGB: HR = 0.68 [95% CI: 0.50, 0.92], *p* = 0.01; RYGB: HR = 0.71 [95% CI: 0.55, 0.91], *p* = 0.01). No other covariate emerged as a significant predictor (Table 3).

## Discussion

In this study, we examined whether preoperative family support predicted long-term outcomes after bariatric surgery. Prior to this research, only a handful of studies had examined this question and were limited in: (i) definitions of family support used (looking at either structural or functional support alone, and either spouses or the family unit), (ii) outcomes measured (focusing only on weight loss), and (iii) time period studied (examining only the short-term window after surgery). Correspondingly, our study was designed to address these gaps.

When we included multiple metrics of family support, marital satisfaction emerged as the primary predictor of weight trajectory. This finding is notable for several reasons. First, two previous studies had observed that marital satisfaction predicted weight loss 6 and 12 months following surgery (17, 18). Extending these studies, our research underscores how satisfaction continues to predict outcomes in the longer term – up to 5 years post-surgery. Second, by assessing both structural and functional support, we resolved a discrepancy in the literature: while several studies had found marital status to confer risk of weight gain, other studies identified marriage as a protective factor (8-10). Based on our research, these conflicting findings may have arisen because marriages differ in quality. This may suggest that it is insufficient to account for structure alone (that is, marital status; 8-10). Instead, the degree to which a marriage is well-functioning may better predict post-surgical outcomes – in accordance with findings of increased divorce rates following bariatric surgery (20, 21).

More broadly, our findings suggest that pre-surgical counselling may benefit from asking patients about spousal support. This aligns with a large body of research describing how family support is critical for a person’s physical health (3, 22, 23). In the case of bariatric surgery, asking patients about their families may guide clinical decisions about patient suitability, or identify subgroups that may require additional post-surgical support. Moving forward, future studies can also explore whether these associations are causal, investigating – for example – whether relationship therapy can boost postoperative weight loss.

While we documented a link between family support and weight trajectories, we did not find any significant association between support and T2DM control. A recent review reported that clinical variables (e.g., use of medication, duration of diabetes) explain the bulk of individual differences relating to postoperative T2DM remission (24). Consequently, psychosocial variables such as family support may have less to contribute within models predicting T2DM outcomes. Further research is needed to explore this possibility.

Several study limitations should be noted. First, family influence is complex, and there are multiple aspects that we were unable to control for. For example, we only examined preoperative family support. Although this strategy allows clinicians to forecast post-surgical weight trajectories, family support is dynamic and may change following surgery (20, 21). Similarly, we did not factor family members’ weight status and weight loss journeys (e.g., whether they had undergone bariatric surgery), nor considered support from persons outside the nuclear family. Further research is thus needed to understand how these nuanced aspects of family support may relate to long-term weight loss outcomes. On a separate note, we also relied on patients’ self-disclosure of family support. Self-reports may be vulnerable to memory or response biases, and future studies would profit from having objective metrics of support (e.g., whether family members accompany the patient at appointments).

## Conclusion

Notwithstanding these limitations, we present the most comprehensive study tracking how family support relates to five-year outcomes after bariatric surgery. Patients who were satisfied with their marriages showed more sustained weight loss over time, alluding to the importance of family involvement in ongoing patient care.

## Data Availability

Data available on request from the authors

## Statement of Human Rights

This study has been approved by the institutional review board (IRB). As a retrospective study, formal consent was not required.

## Acknowledgements

The authors gratefully acknowledge Wong Zhenglong, Jannell Natasha Job, and Kylie Heng for their assistance in data and manuscript preparation.

